# Exploring the causal effect of placental physiology in susceptibility to mental and addictive disorders: a Mendelian randomization study

**DOI:** 10.1101/2024.04.11.24305657

**Authors:** Pablo Jácome-Ferrer, Javier Costas

## Abstract

**Background:** Epidemiological studies have linked low birth weight to psychiatric disorders, including substance use disorders. Genomic analyses suggest a role of placental physiology on psychiatric risk. We investigated whether this association is causally related to impaired trophoblast function.

**Methods:** We conducted a two-sample summary-data Mendelian randomization study using as instrumental variables genetic variants strongly associated with birth weight, whose effect is exerted through the fetal genome, and are located near genes with differential expression in trophoblasts. Eight psychiatric and substance use disorders with >10,000 samples were included as outcomes. The inverse variance weighted method was used as the main analysis and several sensitivity analyses were performed for those significant results.

**Results:** The inverse variance weighted estimate, based on 14 instrumental variables, revealed an association, after correction for multiple tests, between birth weight and broadly-defined depression (beta=-0.165, 95% CI=-0.282 to -0.047, P=0.0059). Sensitivity analyses revealed absence of heterogeneity in the effect of instrumental variables, confirmed by leave-one-out analysis, MR_Egger intercept and MR_PRESSO. The effect was consistent using robust methods. Reverse causality was not detected. The effect was specifically linked to genetic variants near genes involved in trophoblast physiology instead of genes with fetal effect on birth weight or involved in placenta development.

**Conclusion:** Impaired trophoblast functioning, probably leading to reduced fetal brain oxygen and nutrient supply, is causally related to broadly-defined depression. Considering the therapeutic potential of some agents to treat fetal growth restriction, further research on the effect of trophoblast physiology on mental disorders may have future implications in prevention.

## 1 Introduction

Epidemiological data revealed an association between prenatal/perinatal problems, such as gestational diabetes, gestational hypertension, maternal infections during gestation, nutritional deficits during gestation, or preeclampsia and mental disorders (1–3). These agrees with the fetal origins of mental health, framed within the developmental origins of health and disease hypothesis (4). The hypothesis proposes that inappropriate fetal environment may affect brain development, leading to an increased susceptibility to mental disorders later in life.

Low birth weight is commonly used as an easy measure of putative fetal adversity. Meta-analyses of observational studies identified that low birth weight is associated with several mental disorders, such as depression (5), psychosis (2) or autism (3). Large epidemiological studies also reported an association between low birth weight, or small for gestational age, and substance use disorder in adolescence/young adulthood (6) and adults (7,8). Remarkably, Pettersson et al. (8) have also found an association between low birth weight and a latent variable measuring a general factor of psychopathology.

However, association does not imply causality. Mendelian randomization (MR) is a methodological approach to test for causality using genetic predisposition as a proxy for the exposure factor. (9). Arafat and Minica (10) performed a two-sample MR study using GWAS for birth weight from the Early Growth Genetics Consortium (EGGC) (11), as a proxy of exposure to fetal adversity, to test the impact of this exposure on mental disorders, specifically, depression, schizophrenia (SCZ), and attention-deficit/hyperactivity disorder (ADHD). They did not find any evidence of causality. Conversely, Orri et al. (12) revealed a putative causal effect of low birth weight on ADHD by MR. This causal effect was not detected for other mental disorders, such as bipolar disorder (BIP), SCZ, or depression. One explanation for the contradictory results at ADHD may be that while Arafat and Minica (10) considered all SNPs associated with birth weight, Orri et al. (12) limited the selection to those SNPs with direct fetal effect on birth weight, removing those with indirect maternal effect.

Although easy to measure, birth weight has limitations as a proxy for fetal adversity. It does not distinguish between normal growth in constitutionally small but healthy newborns and fetal growth restriction, the condition in which a fetus does not reach its growth potential (13,14). A key player in fetal growth restriction is the placenta. The main cell type of placenta is the trophoblast, whose origin is fetal. Villous cytotrophoblasts are stem cells that give rise to syncytiotrophoblast and extravillous trophoblasts. Syncytiotrophoblasts are involved in most functions of the placenta, such as active transport of nutrients, waste excretion, oxygenation, hormone production, and protection against xenobiotics and the maternal immune system. Extravillous trophoblasts are involved in the invasion of maternal decidua to establish the maternal-fetal circulation through vascular remodeling (15–17).

A role of placenta on risk to psychiatric disorders has been suggested by an interaction between polygenic risk scores (PRS) of SCZ and obstetric complications in the onset of SCZ (18). Furthermore, this interaction was mainly due to PRS of genes highly expressed in placenta and differentially expressed in placenta of complicated pregnancies, referred to as PlacPRS. The interaction, restricted to birth asphyxia, has been confirmed in a Norwegian sample (19). However, it was not replicated in a larger independent study (20). The lack of replication may be due to a type I error or to the use of different scales to measure obstetric complications at each study (12,21). PlacPRS was also negatively associated with neonatal brain volume and cognitive development at 1 year (22). Interestingly, the Norwegian study also detected an interaction between PlacPRS and birth asphyxia in neonatal head circumference (19).

Here, we perform a MR study of birth weight on mental disorders based on a larger GWAS than previous studies (23). Furthermore, we selected as instrumental variables (IVs) those SNPs with fetal effect and close to genes involved in trophoblast biology based on single-cell RNA sequencing studies of the decidual-placental interface (24,25) to distinguish fetal adversity from constitutionally small newborns.

## 2 Materials and Methods

This study was conducted in accordance with the STROBE-MR (Strengthening the Reporting of Observational Studies in Epidemiology-Mendelian Randomization) guidelines for reporting MR studies (Supplementary Table 1) (26).

### 2.1 Selection of genetic instrumental variables

For a SNP to be a valid IV, it must meet the following conditions (referred to as core MR assumptions): it should be associated with the exposure (relevance assumption), not associated with confounders (independence assumption) and only related to the outcome through the exposure (exclusion restriction assumption) (27). SNPs used as IVs were taken from the currently largest GWAS meta-analysis on birth weight (N = 423,683, including samples from EGGC, UK Biobank, and the Icelandic birth register). Birth weight was normalized to a standard normal distribution using rank-based inverse normal transformation prior to analysis (23). The study included information on whether the variants exert their effect directly through the fetal genome or indirectly through the maternal genome, based on a subset of 104,920 Icelandic parent–offspring trios. The selected SNPs accomplished these conditions: i) P < 5x10-8 in the main GWAS of birth weight, i.e., genome-wide significant (GWS) SNPs ii) classified as acting through the fetal effect, and iii) located near genes involved in trophoblast biology. Specifically, all annotated coding genes in the 300 kb region centered on each SNP were collected using the MAGMA (28) auxiliary file for GRCh38 version and the R package “GenomicRanges 1.52.1” (29). Then, the PlacentalCellEnrich tool (30) was used to test for cell-specific expression of these genes in any of the three types of trophoblast cells identified in single-cell RNA sequencing of first trimester human placenta, i.e., villous cytotrophoblast, syncitiotrophoblast, and extravillous trophoblast (24,25). All type of cell-specific expression defined by PlacentaCellEnrich were considering, corresponding to at least five-fold higher expression level in a particular cell type or group of cell types compared to all other cells, or to average levels in all other cells. The default expression threshold of 1 was used. PlacentalCellEnrich was also used to test for cell-specific gene enrichment of the subset of genes around the GWS SNPs with fetal effects, using the hypergeometric test.

Additional IVs were selected in sensitivity analyses. Specifically, all human genes with direct or indirect relationships with the Gene Ontology (GO) (31,32) term GO:0001892, embryonic placenta development, were retrieved using AmiGO 2 (33). SNPs were selected if present a P < 5x10^-8^ in the GWAS of birth weight and are at less than 150 kb of any gene from this list. Another group of SNPs selected as IVs were all independent GWS SNPs with predicted fetal effect by Juliusdottir et al. (23). Finally, subsets of SNPs were selected based on the trophoblast cell type where specific gene expression was detected.

As a measure of the strength of the association between each IV or the overall IV and the exposure, the F-statistic was computed for each SNP using the formula

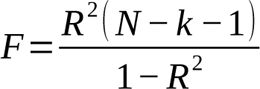

where R^2^ is the proportion of variance explained by each SNP (or their sum, in case of overall IV), N is the sample size and k is equal to 1 when applied to each SNP and equal to the number of IVs when applied to the overall IV. R^2^ was estimated as 2 *β*^2^ *MAF* (1*− MAF*), where β is the beta coefficient and MAF is the minor allele frequency.

### 2.2 Summary data for outcomes

Summary statistics from the largest available GWAS for eight psychiatric disorders with at least 10,000 cases have been selected for this study. The traits included were alcohol dependence (34), ADHD (35), autism spectrum disorder (ASD) (36), BIP (37), cannabis use disorder (CUD) (38), depression (39), post-traumatic stress disorder (PTSD) (40) and SCZ (41) (Supplementary Table 2 lists all GWAS summary statistics used with descriptive information). All included GWAS are from European ancestry samples to ensure similar pattern of linkage disequilibrium.

### 2.3 Mendelian randomization analysis

Two-sample MR analyses were performed between birth weight as exposure and all psychiatric outcomes using the R package “TwoSampleMR 0.5.6” (42). Prior to MR analyses, the exposure and outcome summary statistics were harmonized to ensure that the effects were referenced to the same allele. Palindromic SNPs with minor allele frequency (> 0.4) were removed. Summary statistics of alcohol dependence did not report effect size. In this case, the effect size was estimated from Z score using the following formula

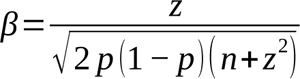

where β is the beta coefficient, z is the Z score, n is the sample size, and p is the effect allele frequency (43). This frequency was taken from the 1000 Genomes Phase 3 European reference panel. In order to estimate the standard errors of beta in ADHD, ASD, PTSD and alcohol dependence summary statistics; the following formula was used

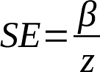

where SE is the standard error of the beta coefficient, β is the beta coefficient and z is the Z score (43).

The causal estimate for each SNP was measured as the ratio between the effect of the SNP on the outcome and the effect of the SNP on the exposure, i.e., the Wald ratio estimate. Wald ratio estimates were meta-analyzed using the random effects inverse-variance weighted method (IVW) as the main MR method to obtain exposure effect size estimates. This is the more commonly used method and is the most powerful method in absence of pleiotropy or if the pleiotropy is balanced (44,45). Heterogeneity was tested using the Q-statistic. Significance for the main results was established as P < 0.00625, corresponding to a Bonferroni’s correction for test on eight outcomes. Considering the existence of genetic correlation between the different outcomes (46), this correction is conservative.

In case of a significant association detected by IVW, several sensitivity tests have been carried out to check the robustness of the results (9,45). Leave-one-out analysis was performed to assess the effect of any single IV on the MR estimate. The MR-Egger regression (47) was used to allow for unbalanced pleiotropic effects. Egger-intercept was used to test for heterogeneity of Wald ratio estimates. MR-PRESSO was used to detect outliers. After outlier removal, MR-PRESSO performs an IVW approach (48). Two methods robust to outliers were used, the weighted median and the weighted mode. The first method is robust to up to 50% of SNPs violating the IV assumptions. The weighted mode is valid under the assumption that the largest subset of IVs with the same estimate are valid instruments, i.e., the majority assumption. This method is sensitive to the bandwidth parameter that defines the clustering of IVs. The default value of 1 was used as first option, while other values were used to analyze robustness of the method. Putative bias due to sample overlap between the exposure and outcome GWAS was quantified using the method proposed by (49). Reverse causality was tested using each psychiatric disorder as exposure and birth weight as outcome.

### 2.4 Ethical considerations

All the data used in this work are de-identified summary statistics data made publicly available after approval by the respective institutional ethical committees of the different consortia. Therefore, no new ethical approval or consent were required.

## 3 Results

### 3.1 Selection of SNPs as instrumental variables based on trophoblast-specific gene expression

There were 351 genes around the 87 lead GWS SNPs, including secondary signals, for fetal growth GWAS with predicted fetal effect. As expected, the most significantly enriched cell types detected by PlacentaCellEnrich corresponded to trophoblast (Supplementary Figure 1). Specifically, syncytiotrophoblasts was significant in the Suryawanshi et al.s’ dataset (24) (adjusted P = 0.047, fold-change = 2.97) and extravillous trophoblasts was significant in the Vento-Tormo et al.s’ dataset (25) (adjusted P = 0.0035, fold-change = 4.82). There were 18 SNPs with a gene specifically-expressed in trophoblast at less than 150 kb. These SNPs were selected as IVs (Table 1). Individual F-statistic ranged from 48.30 to 165.37, indicative of no problems with weak instrument bias. The total number of IVs common to each exposure-outcome analysis was 14-15. The overall F-statistic ranged from 78.21 (depression) to 83.13 (ADHD). The mean F-statistic ranged from 78.63 (depression) to 82.92 (ADHD). The rest of the outcomes employ the same 15 IVs whose overall F-statistic value is 81.78 and mean F-statistic is 81.56. Of the selected IVs, rs7771453, rs577204588, rs1011476 are not found in any outcome GWAS, while rs141845046 and rs10913200 are not found in depression and ADHD, respectively.

**Table 1.**
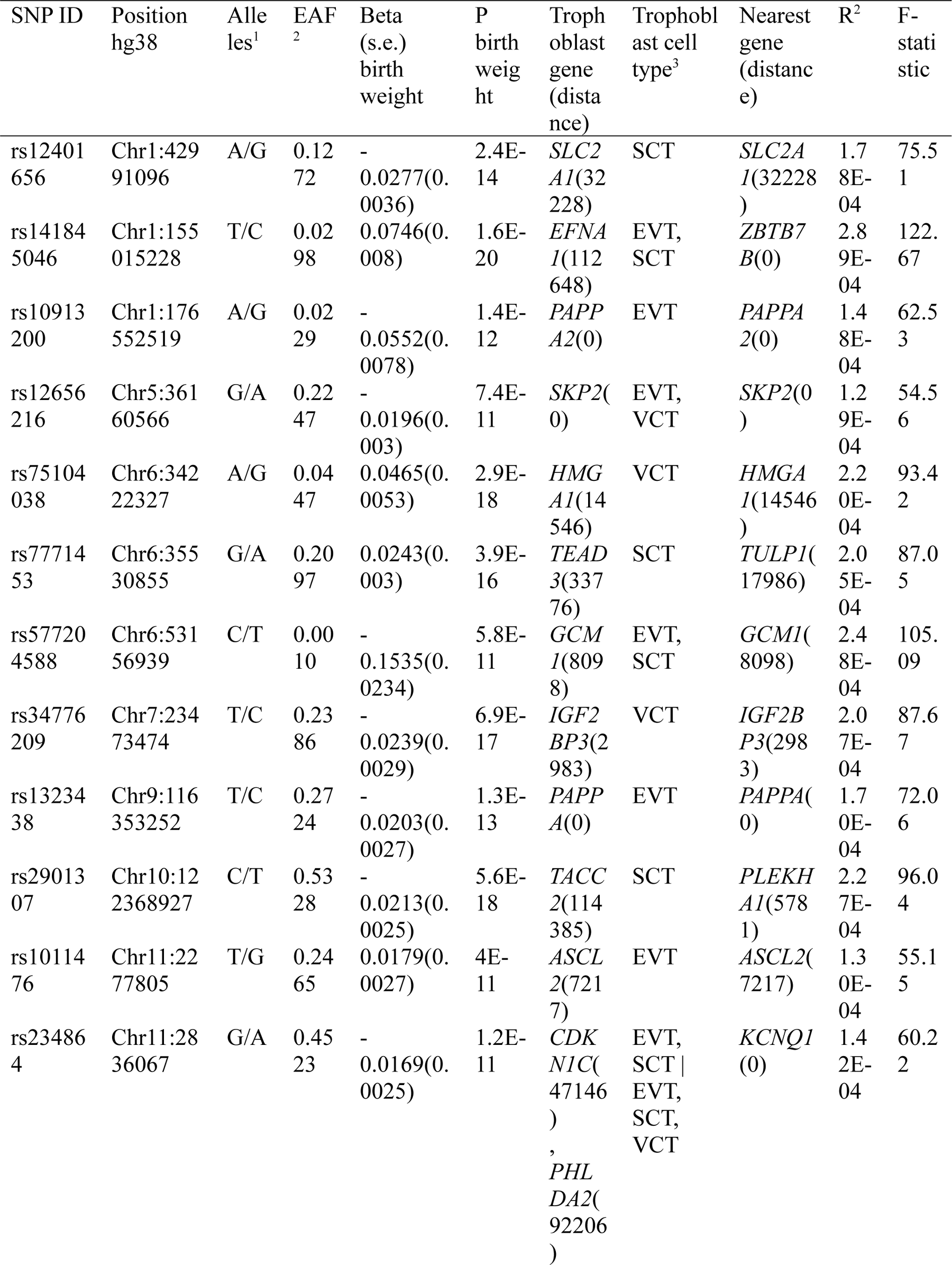
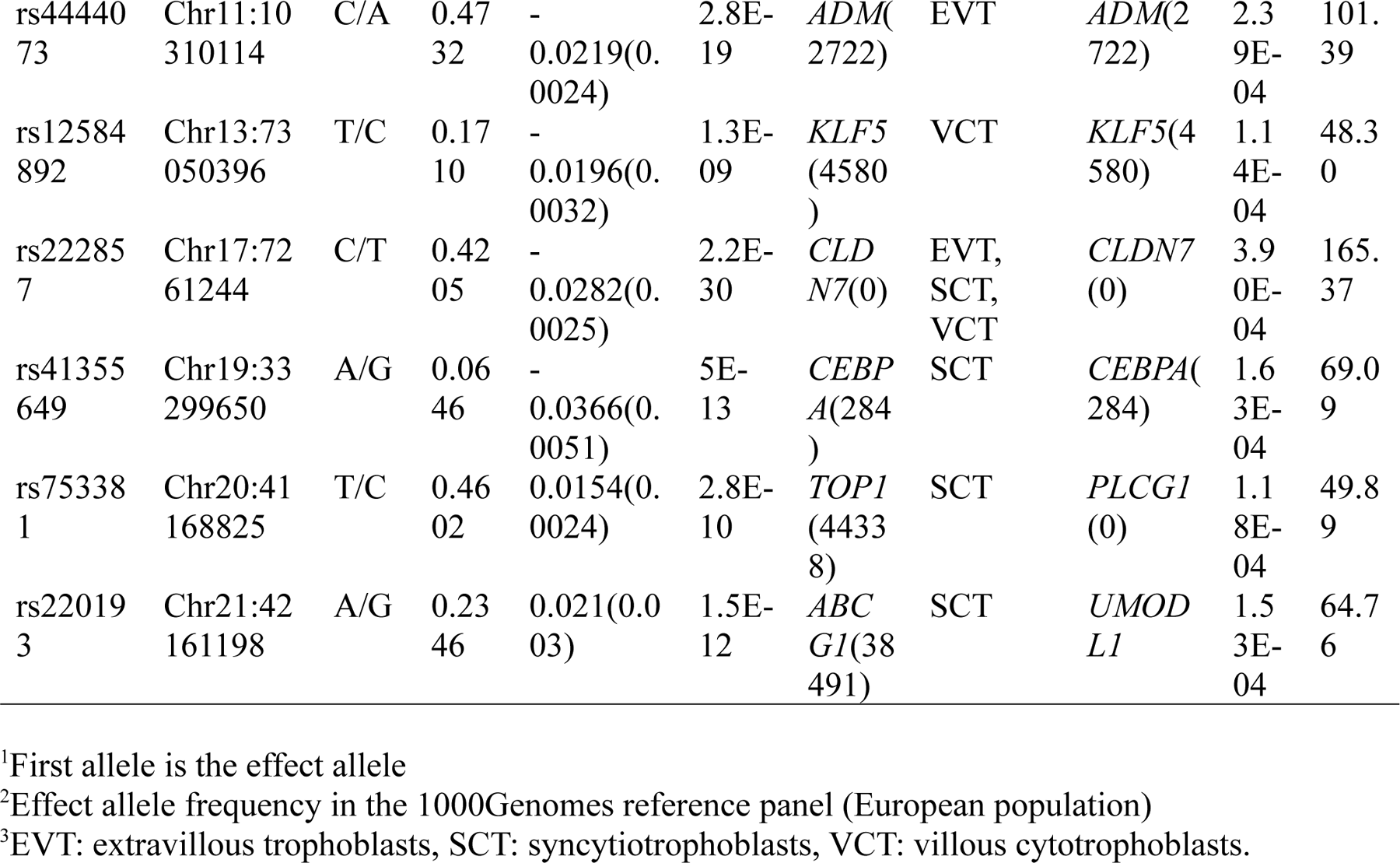
Instrumental Variable (IVs) selected for the Mendelian randomization (MR) analysis.

### 3.2 Main Mendelian randomization analyses

Main results for the MR analyses for each outcome are shown in Figure 1A. IVW method based on 14 trophoblast IVs was significant for depression after correction for multiple tests (beta = -0.165, 95% CI = -0.282 to -0.047, P = 0.0059). The association is in the expected direction, i.e., lower birth weighted is causally associated with increased risk of depression. No other exposure-outcome was significant (P > 0.05 in all cases). When birth weight was considered the outcome and each psychiatric disorder the exposure, there were no association, although PTSD was not tested due to lack of IVs (Figure 1B).

**Figure 1.**
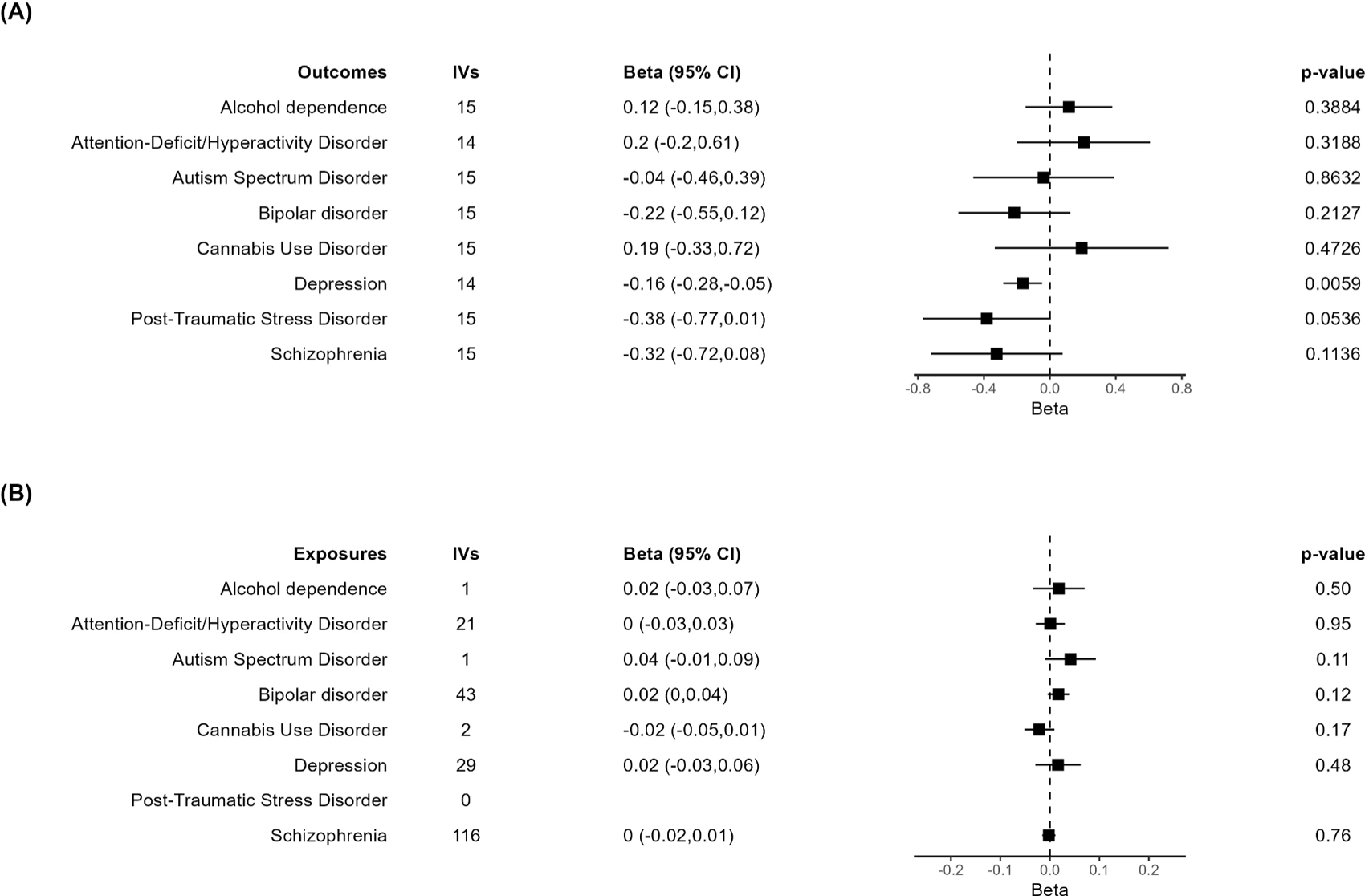
Forest plot of the main results of the MR analysis. (**A**) Results of the MR analysis using the different psychiatric disorders as outcome. (**B**) Results of the MR analysis in which independent SNPs significantly associated with each of the psychiatric disorders were taken as IVs to assess their effect on birth weight, evaluating the inverse causality hypothesis. In case of just one IV, the Wald ratio test is shown. The other results are based on the IVW method.

Assuming that the 217,397 samples from UKBB at the Birth weight GWAS are also among the 361,315 samples from UKBB at the depression GWAS, and that the 11,526 samples from Iceland at the depression GWAS are also among the 125,541 samples from Iceland at the Birth Weight GWAS, the maximum sample overlap is around 45% of the depression samples. Taking into the lower limit of the F-statistic 95% CI, 71.15, the estimated bias associated with overlap was negligible.

### 3.3 Sensitivity analysis for the birth weight – depression association

In agreement with the absence of heterogeneity (Q = 9.07, P = 0.77), the association remained significant in leave-one-out analysis (Figure 2). Furthermore, MR-PRESSO did not detect any outlier using the default outlier significance threshold of 0.05. Egger intercept was not significant (intercept = -0.006, 95% CI = -0.004 to -0.016, P = 0.24), suggesting absence of directional (or unbalanced) pleiotropy. The MR-Egger estimate was not significant (beta = 0.084, 95% CI = -0.324 to 0.493, P = 0.69), and the causal estimate is in the opposite direction (Figure 3). However, there is absence of heterogeneity (Q’ = 7.514, P =0.82), and the difference Q – Q’ was not significant (1.556, P = 0.21), indicating that MR-Egger does not fit substantially better to the data. The weighted median estimate was similar to the IVW (beta = -0.152, 95% CI = -0.313 to 0.009) (Figure 2). The result is near significance (P = 0.0638), in agreement with the low power of the method. Finally, the weighted mode estimate using the default bandwidth of 1 was lower and unsignificant (beta = -0.054, 95% CI = -0.285 to 0.176, P = 0.65). However, the use of different bandwidths gave rise to different conclusions. For instance, the causal estimate using a bandwidth parameter of 2 is more similar to that of the IVW method (beta = -0.13, 95% CI = -0.28 to -0.02, P = 0.089). Using a bandwidth of 2.5 the MR estimate reached significance (beta = -0.139, 95% CI = -0.279 to 0.000, P = 0.050).

**Figure 2.**
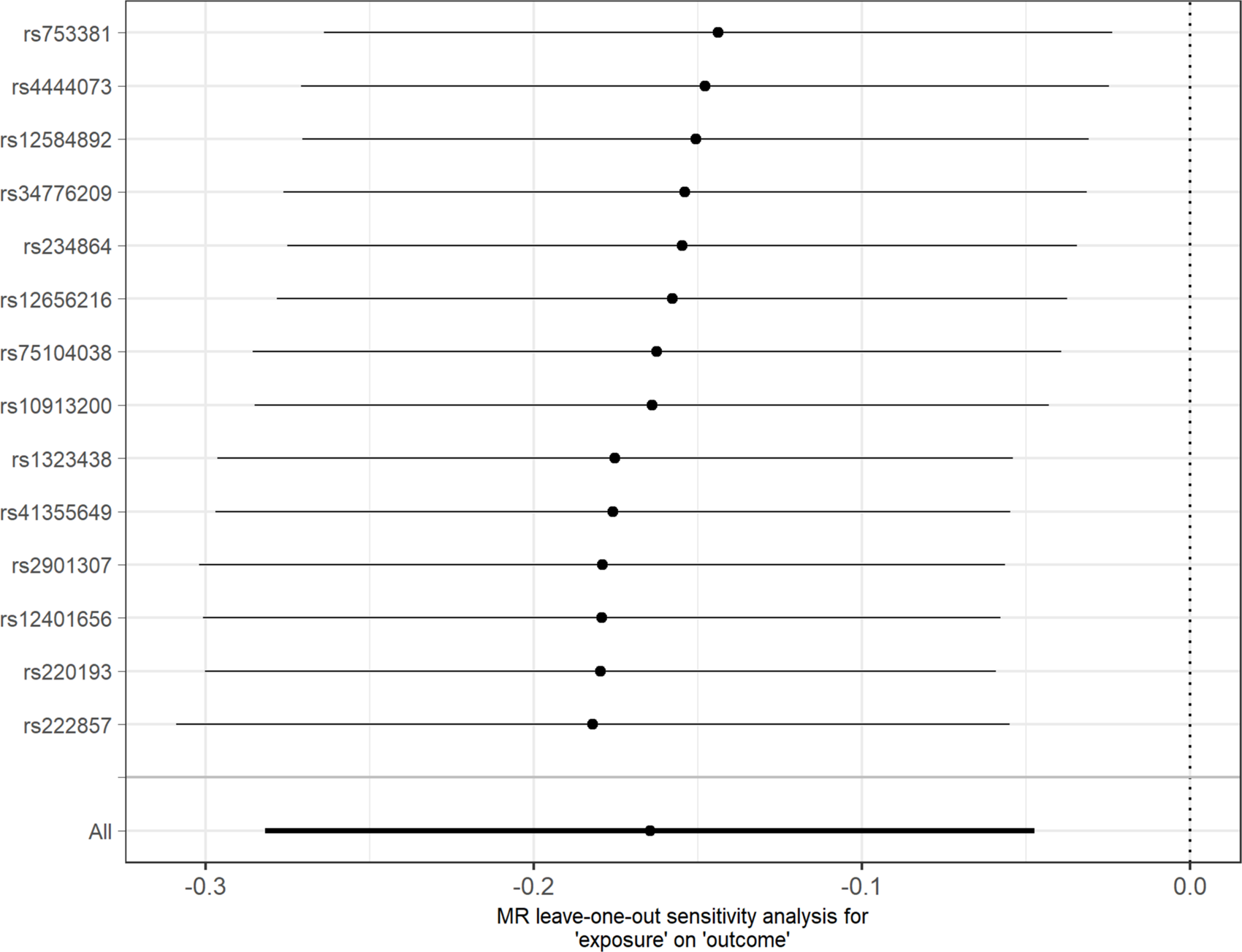
Leave-one-out sensitivity analysis for depression as outcome. The y-axis indicates the SNP that is removed in each analysis. The X-axis shows the beta (dot) and 95% CI (line) for each analysis.

**Figure 3.**
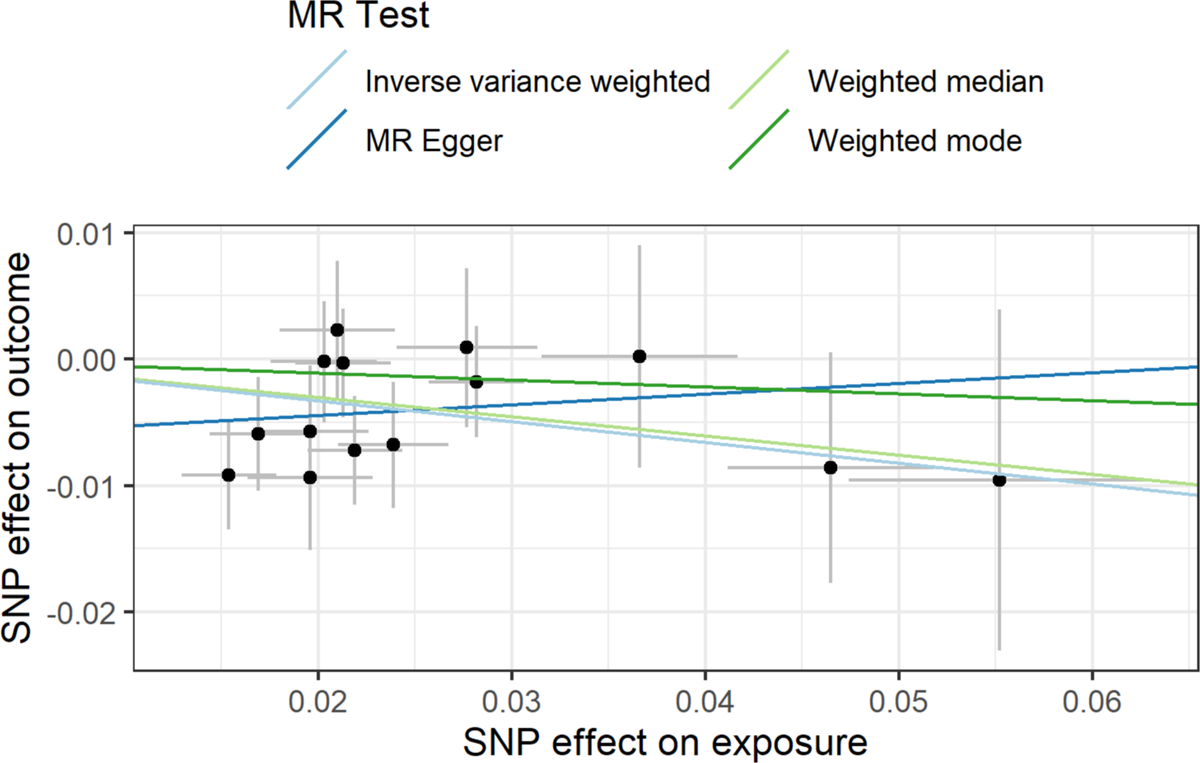
Scatter plot showing the beta effects of SNPs on exposure (birthweight) and outcome (depression). The estimates are represented as dots, with 95% CI represented by horizontal and vertical lines, respectively. The slope of each colored line corresponds to the estimated causal effect by each MR method.

A final set of sensitivity analyses considered distinct IV selection. To test for the effect of trophoblast specific-expression, MR IVW was performed using all independent GWS SNPs for birth weight with predicted fetal effect, n = 79. The causal effect estimate was not significantly different from 0 (beta = -0.009, 95% CI = -0.092 to 0.075, P = 0.84), in agreement with a specific role for trophoblast physiology. Another IV selection included 11 SNPs located near genes involved in embryonic placental development according to GO (Supplementary Table 3). 5 SNPs are common to those from the main analysis based on trophoblast specific-expression. Once again, the result lacked significance (beta = -0.019, 95% CI = -0.175 to 0.136, P = 0.81). Lastly, when the IVs were selected based on the specific trophoblast cell type where specific gene expression was present, IVW estimates were significant in extravillous trophoblasts and villous cytotrophoblasts but not in syncytiotrophoblasts (Figure 4).

**Figure 4.**
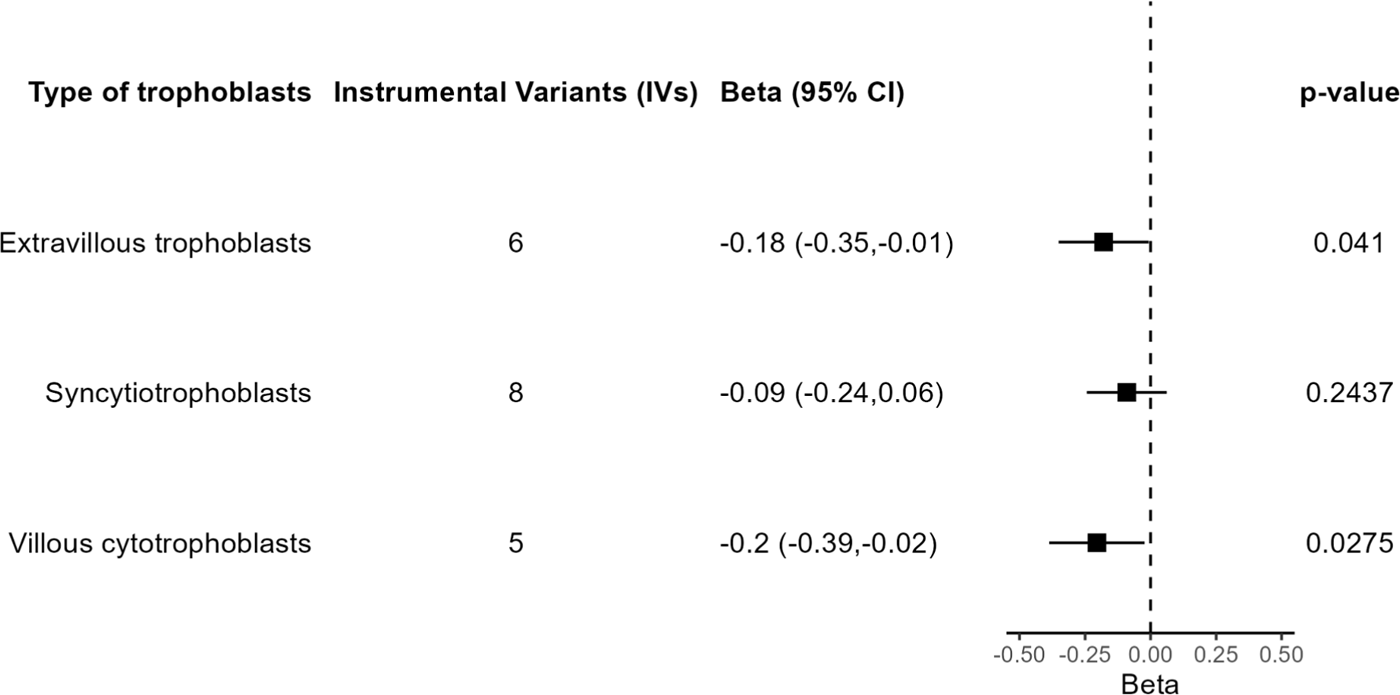
Results of the MR analysis based on subsets of IVs according to trophoblast cell type.

## 4 Discussion

Using a MR approach, we present evidence for the involvement of placenta in susceptibility to broadly defined depression. Specifically, we selected SNPs with fetal effect on birth weight and near genes with differential expression in trophoblasts as IVs for oxygen and nutrient supply to the fetus. However, we did not detect any significant result with other psychiatric or substance use disorders. Reverse causality was also discarded, as none of the sets of IVs chosen for each disorder had a significant effect on birth weight.

A previous MR study analyzed effects of birth weight on several psychiatric disorders, using SNPs with direct fetal effect on birth weight as IVs (12). In contrast with our results, the authors found evidence of an effect of birth weight on ADHD and PTSD but not on depression. These different findings may be related to the selection of IVs. We have chosen SNPs with fetal effect on birth weight and near genes with differential gene expression in trophoblasts as a way to identify IVs specifically involved in oxygen and nutrient supply to the fetus instead of SNPs related to normal growth in constitutionally small but healthy newborns. The relevance of the selection based on differential gene expression in trophoblast was confirmed by lack of significance in sensitivity analyses in depression using as IVs all SNPs with fetal effect on birth weight or SNPs near genes within the GO term “embryonic placenta development”. The use of larger exposure and outcome GWAs in our study may also contribute to differences with the previous one (12). In addition, the estimation of fetal versus maternal effect was based on structural equation modelling in the birth weight GWAS (11) used as exposure by Orri et al. (12) versus phased haplotype data from parent-offspring trios in the one used by us (23).

The outcome GWAS used for depression was based on a broad, highly heterogeneous, phenotype, which includes from people with self-reported depressive symptoms to patients ascertained with a diagnostic interview (39). The genetic susceptibility associated with minimal phenotyping definitions of depression is less specific to depression (50). Thus, it may be possible that the causal association we detected is not specific to depression, but it was the only significant result in MR due to higher power. This agrees with the epidemiological study of (8), who found an association of birth weight with a general psychopathology factor.

The causal effect of trophoblast functioning on susceptibility to psychopathology may be related to the process of invasion of the uterine walls that takes place in the early stages of placentation. The aim of this process is the remodeling of the uterine arteries to provide the fetus a sufficient and permanent supply of oxygen and nutrient (15,16). A problem in the migration capacity of the trophoblasts could potentially result in a reduced number of invaded blood vessels leading to an incomplete remodeling process of the uterine arteries. The reduced blood flow to the fetus will cause a lower or intermittent provision of nutrients and oxygen, and there could be episodes of oxidative stress that can lead to a misfolded protein accumulation (51,52). Hypoxic conditions have also been linked to a reduced activity of differentiation pathways of trophoblasts, such as syncytialization (53,54). Thus, a maternal-fetal interface imbalance mediated by impaired trophoblast function may compromise the normal development of the large human fetal brain, taking into account its high demand of oxygen and nutrients (15,55). Trophoblast invasiveness has recently been suggested as a candidate mechanism to explain the interaction of PlacPRS and obstetric complications on early neurodevelopmental impairment (56).

The findings in this work have to be interpreted in the context of a MR approach. The reliability of MR results is based on several IV assumptions. While the strength of the association between IVs and exposure was confirmed by the F-statistics, other assumptions cannot be formally tested (9). Several sensitivity analyses, such use of methods robust to outliers or analysis of heterogeneity in IVs effects were used to deal with putative horizontal pleiotropy. In addition, we selected IVs near genes with differential expression in trophoblast, as this is the cell type with a more important role in establishment and maintenance of a maternal-fetal interface. Several of the selected genes, such as *ADM*, *ASCL2*, *CDKN1C*, *GCM1*, *HMGA1*, *IGF2BP3*, *PAPPA*, *PAPPA2*, and *PHLDA2*, play a known role in trophoblast physiology and related pathologies, such as intrauterine growth restriction (IUGR) or preeclampsia (57–64). However, we cannot discard an effect of the selected IVs by another mechanism not related with fetal growth restriction.

Another limitation was the low sample size of some of the GWAS of psychiatric disorders reducing the power of the analysis. This is especially problematic in case of substance use disorders. This low power may also be the reason for non-significance of the weighted median analysis in spite of similar effect than the IVW method. Increase of knowledge of trophoblast physiology in near future may lead to identification of more IVs as a way to increase power in addition to increase sample size in GWAS. Finally, the studies included in the PlacentalCellEnrich database (30) use expression data for placentas in the first three months of pregnancy. Although this period is key for establishment of the maternal-fetal interface, it would be necessary to include results from placentas in the second and third trimester to obtain a more complete view of placenta-associated variants.

In summary, we have identified a causal effect of placental impairment in broadly defined depression, probably reflecting an unspecific psychopathology. This adds new data on the current debate about placental role in mental health (21,56,65,66). Although no treatments for fetal growth restriction are currently available, there are several promising agents on clinical trials or preclinical research, acting on processes such as oxygen supply by angiogenesis or vasodilatation, or mitigation of oxidative stress (67,68). Therefore, further research is needed to confirm this causal effect, as it could have clear implications in prevention of mental disorders in near future.

## 6 Author Contributions

JC conceptualized the study and supervised the methodology. PJ-F performed the formal analyses. JC and PJ-F wrote the manuscript. All authors approved the final version of the manuscript.

## 8 Acknowledgments

The authors thanks participants and investigators at deCODE genetics and the Psychiatric Genomics Consortium for providing the summary statistics for this analysis.

## 10 Data Availability Statement

The GWAS summary statistics used for this study can be found in the deCODE genetics (https://www.decode.com/summarydata/) and the Psychiatric Genomics Consortium (https://pgc.unc.edu/for-researchers/download-results/)

## References

1. Arango C, Dragioti E, Solmi M, Cortese S, Domschke K, Murray RM, Jones PB, Uher R, Carvalho AF, Reichenberg A, et al. Risk and protective factors for mental disorders beyond genetics: an evidence-based atlas. World Psychiatry (2021) 20:417–436. doi: 10.1002/WPS.20894

2. Davies C, Segre G, Estradé A, Radua J, De Micheli A, Provenzani U, Oliver D, Salazar de Pablo G, Ramella-Cravaro V, Besozzi M, et al. Prenatal and perinatal risk and protective factors for psychosis: a systematic review and meta-analysis. Lancet Psychiatry (2020) 7:399–410. doi: 10.1016/S2215-0366(20)30057-2

3. Wang C, Geng H, Liu W, Zhang G. Prenatal, perinatal, and postnatal factors associated with autism: A meta-analysis. Medicine (2017) 96: doi: 10.1097/MD.0000000000006696

4. Schlotz W, Phillips DIW. Fetal origins of mental health: Evidence and mechanisms. Brain Behav Immun (2009) 23:905–916. doi: 10.1016/J.BBI.2009.02.001

5. Loret De Mola C, De França GVA, De Avila Quevedo L, Horta BL. Low birth weight, preterm birth and small for gestational age association with adult depression: systematic review and meta-analysis. Br J Psychiatry (2014) 205:340–347. doi: 10.1192/BJP.BP.113.139014

6. Monfils Gustafsson W, Josefsson A, Ekholm Selling K, Sydsjö G. Preterm birth or foetal growth impairment and psychiatric hospitalization in adolescence and early adulthood in a Swedish population-based birth cohort. Acta Psychiatr Scand (2009) 119:54–61. doi: 10.1111/j.1600-0447.2008.01267.x

7. Lahti M, Eriksson JG, Heinonen K, Kajantie E, Lahti J, Wahlbeck K, Tuovinen S, Pesonen AK, Mikkonen M, Osmond C, et al. Late preterm birth, post-term birth, and abnormal fetal growth as risk factors for severe mental disorders from early to late adulthood. Psychol Med (2015) 45:985– 999. doi: 10.1017/S0033291714001998

8. Pettersson E, Larsson H, D’Onofrio B, Almqvist C, Lichtenstein P. Association of Fetal Growth With General and Specific Mental Health Conditions. JAMA Psychiatry (2019) 76:536. doi: 10.1001/JAMAPSYCHIATRY.2018.4342

9. Sanderson E, Glymour MM, Holmes M V., Kang H, Morrison J, Munafò MR, Palmer T, Schooling CM, Wallace C, Zhao Q, et al. Mendelian randomization. Nature Reviews Methods Primers 2022 2:1 (2022) 2:1–21. doi: 10.1038/s43586-021-00092-5

10. Arafat S, Minica CC. Fetal origins of mental disorders? an answer based on mendelian randomization. Twin Research and Human Genetics (2018) 21:485–494. doi: 10.1017/thg.2018.65

11. Warrington NM, Beaumont RN, Horikoshi M, Day FR, Helgeland Ø, Laurin C, Bacelis J, Peng S, Hao K, Feenstra B, et al. Maternal and fetal genetic effects on birth weight and their relevance to cardio-metabolic risk factors. Nature Genetics 2019 51:5 (2019) 51:804–814. doi: 10.1038/s41588-019-0403-1

12. Orri M, Pingault JB, Turecki G, Nuyt AM, Tremblay RE, Côté SM, Geoffroy MC. Contribution of birth weight to mental health, cognitive and socioeconomic outcomes: two-sample Mendelian randomisation. Br J Psychiatry (2021) 219:507–514. doi: 10.1192/BJP.2021.15

13. Kamphof HD, Posthuma S, Gordijn SJ, Ganzevoort W. Fetal Growth Restriction: Mechanisms, Epidemiology, and Management. Maternal-Fetal Medicine (2022) 4:186–196. doi: 10.1097/FM9.0000000000000161

14. Zhang J, Merialdi M, Platt LD, Kramer MS. Defining normal and abnormal fetal growth: promises and challenges. Am J Obstet Gynecol (2010) 202:522–528. doi: 10.1016/J.AJOG.2009.10.889

15. Burton GJ, Fowden AL. The placenta: a multifaceted, transient organ. Philos Trans R Soc Lond B Biol Sci (2015) 370: doi: 10.1098/RSTB.2014.0066

16. Staud F, Karahoda R. Trophoblast: The central unit of fetal growth, protection and programming. Int J Biochem Cell Biol (2018) 105:35–40. doi: 10.1016/J.BIOCEL.2018.09.016

17. Turco MY, Moffett A. Development of the human placenta. Development (2019) 146: doi: 10.1242/DEV.163428

18. Ursini G, Punzi G, Chen Q, Marenco S, Robinson JF, Porcelli A, Hamilton EG, Mitjans M, Maddalena G, Begemann M, et al. Convergence of placenta biology and genetic risk for schizophrenia. Nature Medicine 2018 24:6 (2018) 24:792–801. doi: 10.1038/s41591-018-0021-y

19. Wortinger LA, Shadrin AA, Szabo A, Nerland S, Smelror RE, Jørgensen KN, Barth C, Andreou D, Thoresen M, Andreassen OA, et al. The impact of placental genomic risk for schizophrenia and birth asphyxia on brain development. Translational Psychiatry 2023 13:1 (2023) 13:1–9. doi: 10.1038/s41398-023-02639-4

20. Vassos E, Kou J, Tosato S, Maxwell J, Dennison CA, Legge SE, Walters JTR, Owen MJ, O’Donovan MC, Breen G, et al. Lack of Support for the Genes by Early Environment Interaction Hypothesis in the Pathogenesis of Schizophrenia. Schizophr Bull (2022) 48:20–26. doi: 10.1093/schbul/sbab052

21. Valli I, Gonzalez Segura A, Verdolini N, Garcia-Rizo C, Berge D, Baeza I, Cuesta MJ, Gonzalez-Pinto A, Lobo A, Martinez-Aran A, et al. Obstetric complications and genetic risk for schizophrenia: Differential role of antenatal and perinatal events in first episode psychosis. Acta Psychiatr Scand (2023) doi: 10.1111/acps.13546

22. Ursini G, Punzi G, Langworthy BW, Chen Q, Xia K, Cornea EA, Goldman BD, Styner MA, Knickmeyer RC, Gilmore JH, et al. Placental genomic risk scores and early neurodevelopmental outcomes. Proc Natl Acad Sci U S A (2021) 118: doi: 10.1073/PNAS.2019789118/-/DCSUPPLEMENTAL

23. Juliusdottir T, Steinthorsdottir V, Stefansdottir L, Sveinbjornsson G, Ivarsdottir E V., Thorolfsdottir RB, Sigurdsson JK, Tragante V, Hjorleifsson KE, Helgadottir A, et al. Distinction between the effects of parental and fetal genomes on fetal growth. Nature Genetics 2021 53:8 (2021) 53:1135–1142. doi: 10.1038/s41588-021-00896-x

24. Suryawanshi H, Morozov P, Straus A, Sahasrabudhe N, Max KEA, Garzia A, Kustagi M, Tuschl T, Williams Z. A single-cell survey of the human first-trimester placenta and decidua. Sci Adv (2018) 4: doi: 10.1126/SCIADV.AAU4788

25. Vento-Tormo R, Efremova M, Botting RA, Turco MY, Vento-Tormo M, Meyer KB, Park JE, Stephenson E, Polański K, Goncalves A, et al. Single-cell reconstruction of the early maternal–fetal interface in humans. Nature 2018 563:7731 (2018) 563:347–353. doi: 10.1038/s41586-018-0698-6

26. Skrivankova VW, Richmond RC, Woolf BAR, Yarmolinsky J, Davies NM, Swanson SA, Vanderweele TJ, Higgins JPT, Timpson NJ, Dimou N, et al. Strengthening the Reporting of Observational Studies in Epidemiology Using Mendelian Randomization: The STROBE-MR Statement. JAMA (2021) 326:1614–1621. doi: 10.1001/JAMA.2021.18236

27. Davies NM, Holmes M V., Davey Smith G. Reading Mendelian randomisation studies: a guide, glossary, and checklist for clinicians. BMJ (2018) 362:601. doi: 10.1136/BMJ.K601

28. de Leeuw CA, Mooij JM, Heskes T, Posthuma D. MAGMA: Generalized Gene-Set Analysis of GWAS Data. PLoS Comput Biol (2015) 11:e1004219. doi: 10.1371/JOURNAL.PCBI.1004219

29. Lawrence M, Huber W, Pagès H, Aboyoun P, Carlson M, Gentleman R, Morgan MT, Carey VJ. Software for computing and annotating genomic ranges. PLoS Comput Biol (2013) 9: doi: 10.1371/JOURNAL.PCBI.1003118

30. Jain A, Tuteja G. PlacentaCellEnrich: A tool to characterize gene sets using placenta cell-specific gene enrichment analysis. Placenta (2021) 103:164–171. doi: 10.1016/J.PLACENTA.2020.10.029

31. Ashburner M, Ball CA, Blake JA, Botstein D, Butler H, Cherry JM, Davis AP, Dolinski K, Dwight SS, Eppig JT, et al. Gene Ontology: tool for the unification of biology. Nature Genetics 2000 25:1 (2000) 25:25–29. doi: 10.1038/75556

32. Aleksander SA, Balhoff J, Carbon S, Michael Cherry J, Drabkin HJ, Ebert D, Feuermann M, Gaudet P, Harris NL, Hill DP, et al. The Gene Ontology knowledgebase in 2023. Genetics (2023) 224: doi: 10.1093/genetics/iyad031

33. Carbon S, Ireland A, Mungall CJ, Shu S, Marshall B, Lewis S, Lomax J, Mungall C, Hitz B, Balakrishnan R, et al. AmiGO: online access to ontology and annotation data. Bioinformatics (2009) 25:288–289. doi: 10.1093/BIOINFORMATICS/BTN615

34. Walters RK, Polimanti R, Johnson EC, McClintick JN, Adams MJ, Adkins AE, Aliev F, Bacanu SA, Batzler A, Bertelsen S, et al. Trans-ancestral GWAS of alcohol dependence reveals common genetic underpinnings with psychiatric disorders. Nat Neurosci (2018) 21:1656. doi: 10.1038/S41593-018-0275-1

35. Demontis D, Walters GB, Athanasiadis G, Walters R, Therrien K, Nielsen TT, Farajzadeh L, Voloudakis G, Bendl J, Zeng B, et al. Genome-wide analyses of ADHD identify 27 risk loci, refine the genetic architecture and implicate several cognitive domains. Nature Genetics 2023 55:2 (2023) 55:198–208. doi: 10.1038/s41588-022-01285-8

36. Grove J, Ripke S, Als TD, Mattheisen M, Walters RK, Won H, Pallesen J, Agerbo E, Andreassen OA, Anney R, et al. Identification of common genetic risk variants for autism spectrum disorder. Nat Genet (2019) 51:431–444. doi: 10.1038/S41588-019-0344-8

37. Mullins N, Forstner AJ, O’Connell KS, Coombes B, Coleman JRI, Qiao Z, Als TD, Bigdeli TB, Børte S, Bryois J, et al. Genome-wide association study of more than 40,000 bipolar disorder cases provides new insights into the underlying biology. Nat Genet (2021) 53:817–829. doi: 10.1038/S41588-021-00857-4

38. Johnson EC, Demontis D, Thorgeirsson TE, Walters RK, Polimanti R, Hatoum AS, Sanchez-Roige S, Paul SE, Wendt FR, Clarke TK, et al. A large-scale genome-wide association study meta-analysis of cannabis use disorder. Lancet Psychiatry (2020) 7:1032–1045. doi: 10.1016/S2215-0366(20)30339-4

39. Howard DM, Adams MJ, Clarke TK, Hafferty JD, Gibson J, Shirali M, Coleman JRI, Hagenaars SP, Ward J, Wigmore EM, et al. Genome-wide meta-analysis of depression identifies 102 independent variants and highlights the importance of the prefrontal brain regions. Nature Neuroscience 2019 22:3 (2019) 22:343–352. doi: 10.1038/s41593-018-0326-7

40. Nievergelt CM, Maihofer AX, Klengel T, Atkinson EG, Chen CY, Choi KW, Coleman JRI, Dalvie S, Duncan LE, Gelernter J, et al. International meta-analysis of PTSD genome-wide association studies identifies sex- and ancestry-specific genetic risk loci. Nature Communications 2019 10:1 (2019) 10:1–16. doi: 10.1038/s41467-019-12576-w

41. Trubetskoy V, Pardiñas AF, Qi T, Panagiotaropoulou G, Awasthi S, Bigdeli TB, Bryois J, Chen CY, Dennison CA, Hall LS, et al. Mapping genomic loci implicates genes and synaptic biology in schizophrenia. Nature (2022) 604:502–508. doi: 10.1038/S41586-022-04434-5

42. Hemani G, Zheng J, Elsworth B, Wade KH, Haberland V, Baird D, Laurin C, Burgess S, Bowden J, Langdon R, et al. The MR-Base platform supports systematic causal inference across the human phenome. Elife (2018) 7: doi: 10.7554/ELIFE.34408

43. Zhu Z, Zhang F, Hu H, Bakshi A, Robinson MR, Powell JE, Montgomery GW, Goddard ME, Wray NR, Visscher PM, et al. Integration of summary data from GWAS and eQTL studies predicts complex trait gene targets. Nature Genetics 2016 48:5 (2016) 48:481–487. doi: 10.1038/ng.3538

44. Burgess S, Butterworth A, Thompson SG. Mendelian randomization analysis with multiple genetic variants using summarized data. Genet Epidemiol (2013) 37:658–665. doi: 10.1002/gepi.21758

45. Burgess S, Davey Smith G, Davies NM, Dudbridge F, Gill D, Glymour MM, Hartwig FP, Holmes M V., Minelli C, Relton CL, et al. Guidelines for performing Mendelian randomization investigations. Wellcome Open Res (2020) 4: doi: 10.12688/WELLCOMEOPENRES.15555.2

46. Lee PH, Anttila V, Won H, Feng YCA, Rosenthal J, Zhu Z, Tucker-Drob EM, Nivard MG, Grotzinger AD, Posthuma D, et al. Genomic Relationships, Novel Loci, and Pleiotropic Mechanisms across Eight Psychiatric Disorders. Cell (2019) 179:1469–1482.e11. doi: 10.1016/j.cell.2019.11.020

47. Bowden J, Smith GD, Burgess S. Mendelian randomization with invalid instruments: Effect estimation and bias detection through Egger regression. Int J Epidemiol (2015) 44:512–525. doi: 10.1093/ije/dyv080

48. Verbanck M, Chen CY, Neale B, Do R. Detection of widespread horizontal pleiotropy in causal relationships inferred from Mendelian randomization between complex traits and diseases. Nat Genet (2018) 50:693–698. doi: 10.1038/S41588-018-0099-7

49. Burgess S, Davies NM, Thompson SG. Bias due to participant overlap in two-sample Mendelian randomization. Genet Epidemiol (2016) 40:597–608. doi: 10.1002/GEPI.21998

50. Cai N, Revez JA, Adams MJ, Andlauer TFM, Breen G, Byrne EM, Clarke TK, Forstner AJ, Grabe HJ, Hamilton SP, et al. Minimal phenotyping yields genome-wide association signals of low specificity for major depression. Nat Genet (2020) 52:437–447. doi: 10.1038/S41588-020-0594-5

51. Guzel E, Arlier S, Guzeloglu-Kayisli O, Tabak MS, Ekiz T, Semerci N, Larsen K, Schatz F, Lockwood CJ, Kayisli UA. Endoplasmic Reticulum Stress and Homeostasis in Reproductive Physiology and Pathology. Int J Mol Sci (2017) 18: doi: 10.3390/IJMS18040792

52. Poston L, Raijmakers MTM. Trophoblast Oxidative Stress, Antioxidants and Pregnancy Outcome—A Review. Placenta (2004) 25:S72–S78. doi: 10.1016/J.PLACENTA.2004.01.003

53. Genbacev O, Zhou Y, Ludlow JW, Fisher SJ. Regulation of human placental development by oxygen tension. Science (1997) 277:1669–1672. doi: 10.1126/SCIENCE.277.5332.1669

54. Zhou H, Zhao C, Wang P, Yang W, Zhu H, Zhang S. Regulators involved in trophoblast syncytialization in the placenta of intrauterine growth restriction. Front Endocrinol (Lausanne) (2023) 14:1107182. doi: 10.3389/FENDO.2023.1107182/BIBTEX

55. Moffett A, Loke C. Immunology of placentation in eutherian mammals. Nat Rev Immunol (2006) 6:584–594. doi: 10.1038/NRI1897

56. Ursini G, Di Carlo P, Mukherjee S, Chen Q, Han S, Kim J, Deyssenroth M, Marsit CJ, Chen J, Hao K, et al. Prioritization of potential causative genes for schizophrenia in placenta. Nat Commun (2023) 14: doi: 10.1038/s41467-023-38140-1

57. Dugoff L, Hobbins JC, Malone FD, Porter TF, Luthy D, Comstock CH, Hankins G, Berkowitz RL, Merkatz I, Craigo SD, et al. First-trimester maternal serum PAPP-A and free-beta subunit human chorionic gonadotropin concentrations and nuchal translucency are associated with obstetric complications: A population-based screening study (The FASTER Trial). Am J Obstet Gynecol (2004) 191:1446–1451. doi: 10.1016/j.ajog.2004.06.052

58. Gu C, Park S, Seok J, Jang HY, Bang YJ, Kim GIJ. Altered expression of ADM and ADM2 by hypoxia regulates migration of trophoblast and HLA-G expression†. Biol Reprod (2021) 104:159–169. doi: 10.1093/BIOLRE/IOAA178

59. Jeyarajah MJ, Bhattad GJ, Kelly RD, Baines KJ, Jaremek A, Yang FHP, Okae H, Arima T, Dumeaux V, Renaud SJ. The multifaceted role of GCM1 during trophoblast differentiation in the human placenta. Proc Natl Acad Sci U S A (2022) 119:e2203071119. doi: 10.1073/PNAS.2203071119/SUPPL_FILE/PNAS.2203071119.SD01.TXT

60. Li W, Liu D, Chang W, Lu X, Wang YL, Wang H, Zhu C, Lin HY, Zhang Y, Zhou J, et al. Role of IGF2BP3 in trophoblast cell invasion and migration. Cell Death & Disease 2014 5:1 (2014) 5:e1025–e1025. doi: 10.1038/cddis.2013.545

61. Matsubara K, Matsubara Y, Uchikura Y, Takagi K, Yano A, Sugiyama T. HMGA1 Is a Potential Driver of Preeclampsia Pathogenesis by Interference with Extravillous Trophoblasts Invasion. Biomolecules 2021, Vol 11, Page 822 (2021) 11:822. doi: 10.3390/BIOM11060822

62. Neuman RI, Alblas van der Meer MM, Nieboer D, Saleh L, Verdonk K, Kalra B, Kumar A, Alpadi K, van den Meiracker AH, Visser W, et al. PAPP-A2 and Inhibin A as Novel Predictors for Pregnancy Complications in Women With Suspected or Confirmed Preeclampsia. Journal of the American Heart Association: Cardiovascular and Cerebrovascular Disease (2020) 9: doi: 10.1161/JAHA.120.018219

63. Simmers MD, Hudson KM, Baptissart M, Cowley M. Epigenetic control of the imprinted growth regulator Cdkn1c in cadmium-induced placental dysfunction. Epigenetics (2022) doi: 10.1080/15592294.2022.2088173

64. Song G, Jin F. RhoGDI1 interacts with PHLDA2, suppresses the proliferation, migration, and invasion of trophoblast cells, and participates in the pathogenesis of preeclampsia. Hum Cell (2022) 35:1440–1452. doi: 10.1007/S13577-022-00746-W

65. Ursini G, Weinberger DR. Replicating G × E: The Devil and the Details. Schizophr Bull (2022) 48:4–4. doi: 10.1093/SCHBUL/SBAB109

66. Vassos E, Murray RM. The Jury Is Still out on Placental Genes and Obstetric Complications. Schizophr Bull (2022) 48:5–5. doi: 10.1093/SCHBUL/SBAB117

67. Colson A, Sonveaux P, Debiève F, Sferruzzi-Perri AN. Adaptations of the human placenta to hypoxia: opportunities for interventions in fetal growth restriction. Hum Reprod Update (2021) 27:531–569. doi: 10.1093/HUMUPD/DMAA053

68. Groom KM, David AL. The role of aspirin, heparin, and other interventions in the prevention and treatment of fetal growth restriction. Am J Obstet Gynecol (2018) 218:S829–S840. doi: 10.1016/J.AJOG.2017.11.565

