## Supplementary Figures and Tables for "Exploring the causal effect of placental physiology in susceptibility to mental and addictive disorders: a Mendelian randomization study"

### *Supplementary Material*

#### 1 Supplementary Figures and Tables

##### 1.1 Supplementary Figures

**Supplementary Figure 1.** Most significantly enriched cell types detected by each dataset in PlacentaCellEnrich (1). Vento-Tormo dataset (2) (**A**) and Suryawanshi dataset (3) (**B**).

**A**

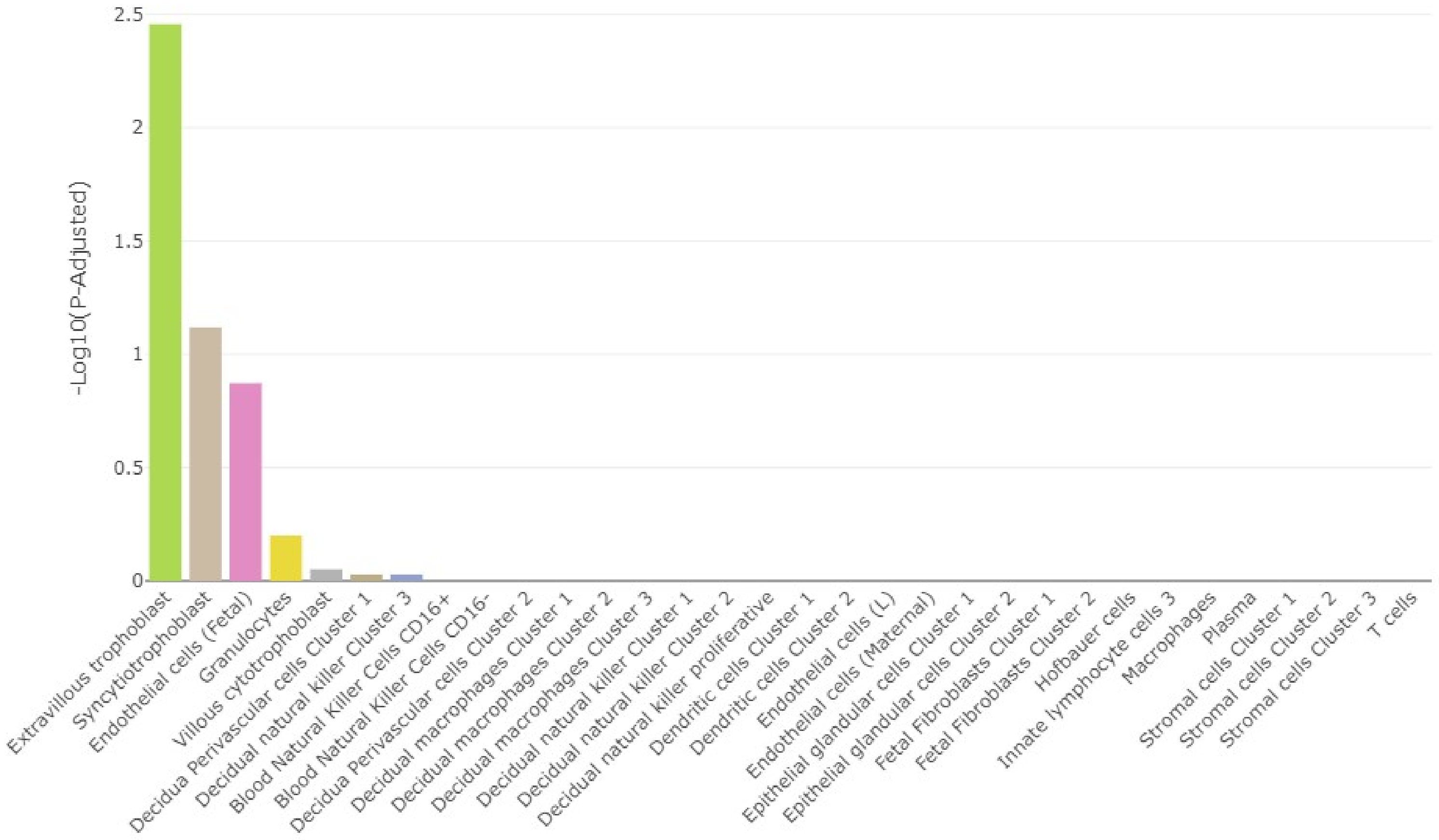

-Log10(P-Adjusted)
2.5
2
1.5
1
0.5
0
Extravillous trophoblast
Syncytiotrophoblast
Endothelial cells (Fetal)
Granulocytes
Villous cytotrophoblast
Decidua Perivascular cells Cluster 1
Decidual natural killer Cluster 3
Blood Natural Killer Cells CD16+
Blood Natural Killer Cells CD16-
Decidua Perivascular cells Cluster 2
Decidual macrophages Cluster 1
Decidual macrophages Cluster 2
Decidual macrophages Cluster 3
Decidual natural killer Cluster 1
Decidual natural killer Cluster 2
Decidual natural killer proliferative
Dendritic cells Cluster 1
Dendritic cells Cluster 2
Endothelial cells (L)
Endothelial cells (Maternal)
Epithelial glandular cells Cluster 1
Epithelial glandular cells Cluster 2
Fetal Fibroblasts Cluster 1
Fetal Fibroblasts Cluster 2
Hofbauer cells
Innate lymphocyte cells 3
Macrophages
Plasma
Stromal cells Cluster 1
Stromal cells Cluster 2
Stromal cells Cluster 3
T cells

B

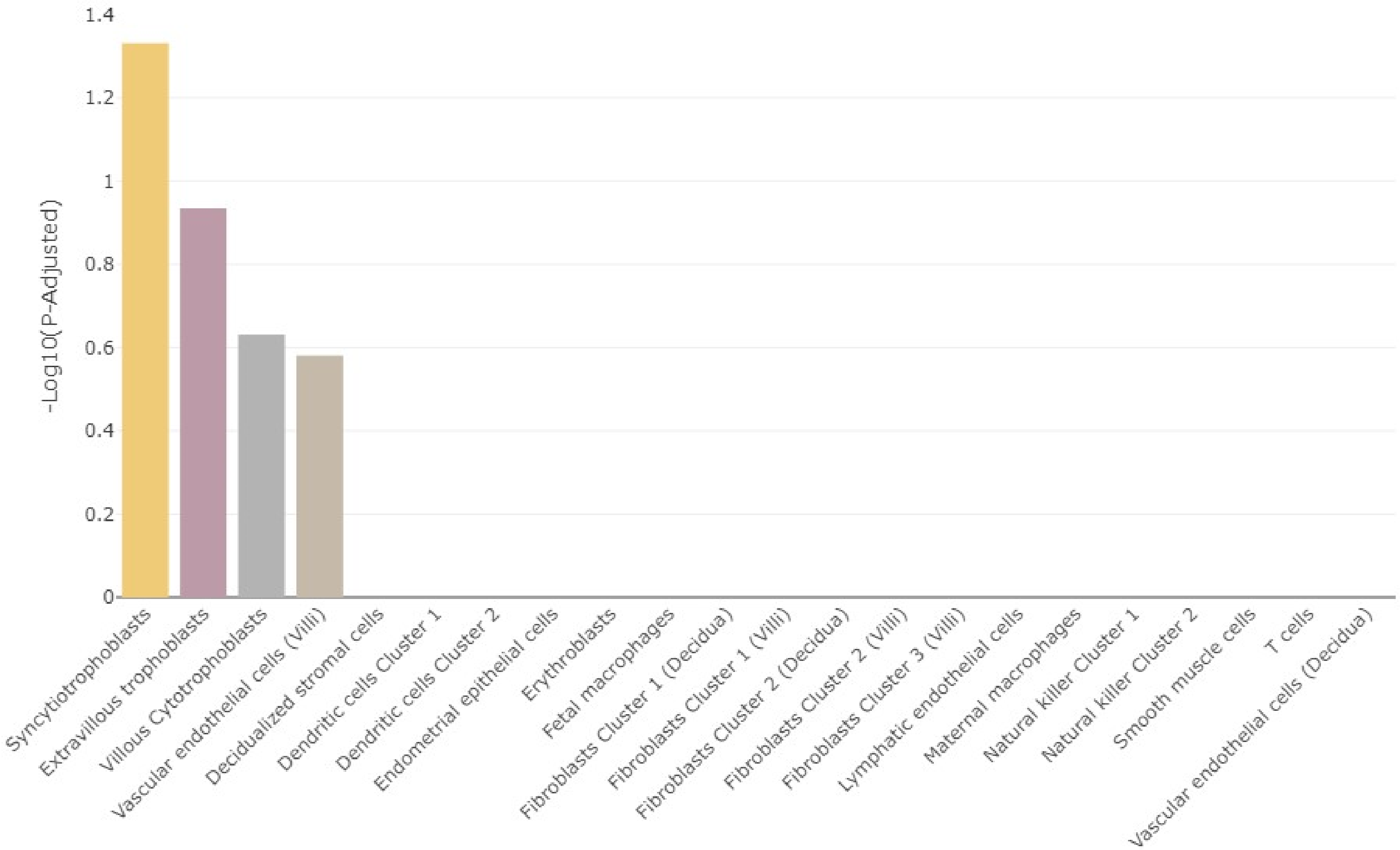
1.4
1.2
1
0.8
0.6
0.4
0.2
0
-Log10(P-Adjusted)
Syncytiotrophoblasts
Extravillous trophoblasts
Villous Cytotrophoblasts
Vascular endothelial cells (Villi)
Decidualized stromal cells
Dendritic cells Cluster 1
Dendritic cells Cluster 2
Endometrial epithelial cells
Erythroblasts
Fetal macrophages
Fibroblasts Cluster 1 (Decidua)
Fibroblasts Cluster 1 (Villi)
Fibroblasts Cluster 2 (Decidua)
Fibroblasts Cluster 2 (Villi)
Fibroblasts Cluster 3 (Villi)
Lymphatic endothelial cells
Maternal macrophages
Natural killer Cluster 1
Natural killer Cluster 2
Smooth muscle cells
T cells
Vascular endothelial cells (Decidua)

#### 1.2 Supplementary Tables

**Supplementary Table 1.** STROBE-MR checklist (4).

| Item No. | Section | Checklist item | Location / relevant text from manuscript |
|---|---|---|---|
| **TITLE AND ABSTRACT** | | | |
| 1 | Title and abstract | Indicate Mendelian randomization (MR) as the study's design in the title and/or the abstract if that is a main purpose of the study | Title: "Role of placental physiology in susceptibility to mental and addictive disorders: a Mendelian randomization study"<br>Abstract: "To test this hypothesis, we conducted a two-sample summary-data Mendelian randomization study using as instrumental variables genetic variants strongly associated with birth weight, whose effect is exerted through the fetal genome, and are located near genes with differential expression in trophoblasts." |
| **INTRODUCTION** | | | |
| 2 | **Background** | Explain the scientific background and rationale for the reported study. What is the exposure? Is a potential causal relationship between exposure and outcome plausible? Justify why MR is a helpful method to address the study question | Introduction, par. 1-3<br>"Epidemiological data revealed an association between prenatal/perinatal problems, such as gestational diabetes, gestational hypertension, maternal infections during gestation, nutritional deficits during gestation, or preeclampsia and mental disorders [2–4]. These agrees with ..." |
| 3 | **Objectives** | State specific objectives clearly, including pre-specified causal hypotheses (if any). State that MR is a method that, under specific assumptions, intends to estimate causal effects | Introduction, par. 3<br>"However, association does not imply causality. Mendelian randomization (MR) is a methodological approach to test for causality using genetic predisposition as a proxy for the exposure factor."<br>Introduction, par. 6:<br>"Confirmation of involvement of placenta in psychiatric risk may have important implications in prevention. Therefore, here we perform a MR study of birth weight on mental disorders based on a larger GWAS than previous studies. We selected as IVs those SNPs with fetal effect and close |

| | | | |
|---|---|---|---|
| | | | to genes involved in trophoblast biology based on single-cell RNA sequencing studies of the decidual-placental interface to distinguish fetal adversity from constitutionally small newborns. " |
| **METHODS** | | | |
| 4 | **Study design and data sources** | Present key elements of the study design early in the article. Consider including a table listing sources of data for all phases of the study. For each data source contributing to the analysis, describe the following: | |
| | a) | Setting: Describe the study design and the underlying population, if possible. Describe the setting, locations, and relevant dates, including periods of recruitment, exposure, follow-up, and data collection, when available. | Materials and Methods, subsection "Selection of genetic instrumental variables", par. 1:<br>"The study meta-analyzed the largest previous GWAS to date from European ancestry samples [14], which included individuals from the EGGC and UK Biobank (UKBB), with newborn data from the Icelandic birth register".<br>Materials and Methods, subsection "Summary data for outcomes":<br>"All included GWAS are from European ancestry samples to ensure similar pattern of linkage disequilibrium."<br>Supplementary Table 2. |
| | b) | Participants: Give the eligibility criteria, and the sources and methods of selection of participants. Report the sample size, and whether any power or sample size calculations were carried out prior to the main analysis | Supplementary Table 2.<br>Power calculations were not carried out, but we have used the largest GWAS to date and consider only those GWAS with large sample size.<br>Materials and Methods, subsection "Summary data for outcomes":<br>"Summary statistics from the largest available GWAS for eight psychiatric disorders with at least 10,000 cases have been selected for this study." |
| | c) | Describe measurement, quality control and selection of genetic variants | Materials and Methods, subsection "Selection of genetic instrumental variables", par. 1 |
| | d) | For each exposure, outcome, and other relevant variables, describe methods of assessment and diagnostic | Supplementary Table 2. |

| | | | |
|---|---|---|---|
| | | criteria for diseases | |
| | e) | Provide details of ethics committee approval and participant informed consent, if relevant | Declarations, subsection "Ethics approval and consent to participate":<br>"All the data used in this work are de-identified summary statistics data made publicly available after approval by the respective institutional ethical committees of the different consortia. Therefore, no new ethical approval or consent were required". |
| 5 | **Assumptions** | Explicitly state the three core IV assumptions for the main analysis (relevance, independence and exclusion restriction) as well assumptions for any additional or sensitivity analysis | Materials and Methods, subsection "Summary data for outcomes":<br>"For a SNP to be a valid IV, it must meet the following conditions (referred to as core MR assumptions): it should be associated with the exposure (relevance assumption), not associated with confounders (independence assumption) and only related to the outcome through the exposure (exclusion restriction assumption)" |
| 6 | **Statistical methods: main analysis** | Describe statistical methods and statistics used | |
| | a) | Describe how quantitative variables were handled in the analyses (i.e., scale, units, model) | Materials and Methods, subsection "Selection of genetic instrumental variables", par. 1:<br>"Birth weight was normalized to a standard normal distribution using rank-based inverse normal transformation prior to analysis". |
| | b) | Describe how genetic variants were handled in the analyses and, if applicable, how their weights were selected | Materials and Methods, subsection "Selection of genetic instrumental variables", par. 1:<br>"The selected SNPs accomplished these conditions: i) P < 5x10-8 in the main GWAS of …"<br>Materials and Methods, subsection "Mendelian randomization analysis", par. 1:<br>"Prior to MR analyses, the exposure and outcome summary statistics were harmonized using "TwoSampleMR" to ensure that the effects were referenced to the same allele. Palindromic SNPs with minor allele frequency (> 0.4) were removed. Summary statistics of alcohol dependence did not report effect size. In this case, the effect size was estimated from ..." |
| | c) | Describe the MR estimator (e.g., two-stage least squares, Wald ratio) and related statistics. Detail the included covariates and, in case of two-sample | Materials and Methods, subsection "Mendelian randomization analysis", par. 2:<br>"The causal estimate for each SNP was measured as the ratio between the effect of the SNP on the outcome and the effect of the SNP on the |

| | | | |
|---|---|---|---|
| | | MR, whether the same covariate set was used for adjustment in the two samples | exposure, i.e., the Wald ratio estimate. ” |
| | d) | Explain how missing data were addressed | Although not explicitly mentioned it, we did not try to infer missing data. The existence of missing data is mentioned in Results, subsection “Selection of SNPs as instrumental variables based on trophoblast-specific gene expression”:<br>“Of the selected IVs, rs7771453, rs577204588, rs1011476 are not found in any outcome GWAS, while rs141845046 and rs10913200 are not found in depression and ADHD, respectively.” |
| | e) | If applicable, indicate how multiple testing was addressed | Materials and Methods, subsection “Mendelian randomization analysis”, par. 2:<br>“Significance for the main results was established as P < 0.00625, corresponding to a Bonferroni’s correction for test on eight outcomes”. |
| 7 | **Assessment of assumptions** | Describe any methods or prior knowledge used to assess the assumptions or justify their validity | Materials and Methods, subsection “Selection of genetic instrumental variables”, par. 1:<br>“The selected SNPs accomplished these conditions: i) P < 5x10-8 in the main GWAS of birth weight, i.e., genome-wide significant (GWS) SNPs ii) classified as acting through the fetal effect, and iii) located near genes involved in trophoblast biology. […] Then, the PlacentalCellEnrich tool [32] was used to test for cell-specific expression of these genes in any of the three types of trophoblast cells identified in single-cell RNA sequencing of first trimester human placenta, i.e., villous cytotrophoblast, syncitiotrophoblast, and extravillous trophoblast [28,29].”<br>Materials and Methods, subsection “Selection of genetic instrumental variables”, par. 3:<br>“As a measure of the strength of the association between each IV or the overall IV and the exposure, the F-statistic was computed for each SNP using the formula” |
| 8 | **Sensitivity analyses and additional analyses** | Describe any sensitivity analyses or additional analyses performed (e.g., comparison of effect estimates from different approaches, independent replication, bias analytic techniques, | Materials and Methods, subsection “Selection of genetic instrumental variables”, par. 2:<br>“Additional IVs were selected in sensitivity analyses. Specifically, all human genes with direct or indirect relationships with...”<br>Materials and Methods, subsection “Mendelian randomization analysis”, |

| | | | |
|---|---|---|---|
| | | validation of instruments, simulations) | par. 3.<br>“In case of a significant association detected by IVW, several sensitivity tests have been carried out to check the robustness of the results ...” |
| 9 | **Software and pre-registration** | | |
| | a) | Name statistical software and package(s), including version and settings used | Materials and Methods, subsection “Mendelian randomization analysis”, par. 1:<br>“Two-sample MR analyzes were performed between birth weight as exposure and all psychiatric outcomes using the R package "TwoSampleMR 0.5.6"<br>Materials and Methods, subsection “Selection of genetic instrumental variables”, par. 3:<br>“This method is sensitive to the bandwidth parameter that defines the clustering of IVs. The default value of 1 was used as first option, while other values were used to analyze robustness of the method. ” |
| | b) | State whether the study protocol and details were pre-registered (as well as when and where) | The study protocol was not pre-registered |
| **RESULTS** | | | |
| 10 | **Descriptive data** | | |
| | a) | Report the numbers of individuals at each stage of included studies and reasons for exclusion. Consider use of a flow diagram | Not applicable to our study, as it is based on summary statistics from previous GWAS. |
| | b) | Report summary statistics for phenotypic exposure(s), outcome(s), and other relevant variables (e.g., means, SDs, proportions) | Supplementary Table 2. |
| | c) | If the data sources include meta-analyses of previous studies, provide the assessments of heterogeneity across these studies | Not applicable |
| | d) | For two-sample MR: | As mentioned in Materials and Methods, all samples are of European |

| | | | |
|---|---|---|---|
| | | i. Provide justification of the similarity of the genetic variant-exposure associations between the exposure and outcome samples | ancestry.<br>Materials and Methods, subsection “Selection of genetic instrumental variables”, par. 2:<br>“The study meta-analyzed the largest previous GWAS to date from European ancestry samples [14], which included individuals from the EGGC and UK Biobank (UKBB), with newborn data from the Icelandic birth register, resulting in a total sample size of 423,683 individuals.”<br>Materials and Methods, subsection “Summary data for outcomes”:<br>“All included GWAS are from European ancestry samples to ensure similar pattern of linkage disequilibrium.” |
| | | ii. Provide information on the number of individuals who overlap between the exposure and outcome studies | It is not possible to know the number of overlapping samples. But we took this into account:<br>Materials and Methods, subsection “Mendelian randomization analysis”, par. 3.:<br>“Putative bias due to sample overlap between the exposure and outcome GWAS was quantified using the method proposed by Burgess et al.”<br>Results, subsection “Main Mendelian randomization analysis”, par. 2:<br>“Assuming that the 217,397 samples from UKBB at the Birth weight GWAS are also among the 361,315 samples from UKBB at the depression GWAS, and that the 11,526 samples from Iceland at the depression GWAS are also among the 125,541 samples from Iceland at the Birth Weight GWAS, the maximum sample overlap is around 45% of the depression samples. Taking into the lower limit of the F-statistic 95% CI, 71.15, the estimated bias associated with overlap was negligible.” |
| 11 | **Main results** | | |
| | a) | Report the associations between genetic variant and exposure, and between genetic variant and outcome, preferably on an interpretable scale | Table 1 (exposure), Fig. 3. |
| | b) | Report MR estimates of the relationship between exposure and outcome, and the measures of uncertainty from the MR analysis, on an interpretable scale, such as odds | Fig. 1A.<br>Results, subsection “Main Mendelian randomization analysis”, par. 1:<br>“IVW method based on 14 trophoblast IVs was significant for depression after correction for multiple tests (beta = -0.165, 95% CI = -0.282 to -0.047, P = 0.0059)”. |

| | | | |
|---|---|---|---|
| | | ratio or relative risk per SD difference | |
| | c) | If relevant, consider translating estimates of relative risk into absolute risk for a meaningful time period | Not applicable |
| | d) | Consider plots to visualize results (e.g., forest plot, scatterplot of associations between genetic variants and outcome versus between genetic variants and exposure) | Fig. 1A, Fig. 3 |
| 12 | **Assessment of assumptions** | | |
| | a) | Report the assessment of the validity of the assumptions | Results Subsection "Selection of SNPs as instrumental variables based on trophoblast-specific gene expression":<br>"Individual F-statistic ranged from 48.30 to 165.37, indicative of no problems with weak instrument bias. The total number of IVs common to each exposure-outcome analysis was 14-15. The overall F-statistic ranged from 78.21 (depression) to 83.13 (ADHD). The mean F-statistic ranged from 78.63 (depression) to 82.92 (ADHD). The rest of the outcomes employ the same 15 IVs whose overall F-statistic value is 81.78 and mean F-statistic is 81.56."<br>Results Subsection "Sensitivity analysis for the birth weight – depression association":<br>"Egger intercept was not significant (intercept = -0.006, 95% CI = -0.004 to -0.016, P = 0.24), suggesting absence of directional (or unbalanced) pleiotropy." |
| | b) | Report any additional statistics (e.g., assessments of heterogeneity across genetic variants, such as $I^2$, Q statistic or E-value) | Results Subsection "Sensitivity analysis for the birth weight – depression association":<br>"In agreement with the absence of heterogeneity (Q = 9.07, P = 0.77), the association remained significant in leave-one-out analysis (Figure 2). Furthermore, MR-PRESSO did not detect any outlier using the default outlier significance threshold of 0.05. […] However, there is absence of heterogeneity (Q' = 7.514, P =0.82), and the difference Q – Q' was not significant (1.556, P = 0.21), indicating that MR-Egger does not fit substantially better to the data." |

| | | | |
|---|---|---|---|
| 13 | **Sensitivity analyses and additional analyses** | | |
| | a) | Report any sensitivity analyses to assess the robustness of the main results to violations of the assumptions | Results Subsection "Sensitivity analysis for the birth weight – depression association"<br>"The MR-Egger estimate was not significant (beta = 0.084, 95% CI = -0.324 to 0.493, P = 0.69), and the causal estimate is in the opposite direction (Figure 3). However, [...], indicating that MR-Egger does not fit substantially better to the data. The weighted median estimate was similar to the IVW (beta = -0.152, 95% CI = -0.313 to 0.009) (Figure 2). The result is near significance (P = 0.0638), in agreement with the low power of the method. Finally, the weighted mode ..." |
| | b) | Report results from other sensitivity analyses or additional analyses | Fig. 2, Fig. 3, Table S2. Results Subsection "Sensitivity analysis for the birth weight – depression association" |
| | c) | Report any assessment of direction of causal relationship (e.g., bidirectional MR) | Fig. 1B.<br>Results, subsection "Main Mendelian randomization analysis", par. 1:<br>"When birth weight was considered the outcome and each psychiatric disorder the exposure, there were no association, although PTSD was not tested due to lack of IVs" |
| | d) | When relevant, report and compare with estimates from non-MR analyses | Not applicable |
| | e) | Consider additional plots to visualize results (e.g., leave-one-out analyses) | Leave-one-out analyses: Fig. 2<br>Scatterplot of SNP effects: Fig. 3 |
| **DISCUSSION** | | | |
| 14 | **Key results** | Summarize key results with reference to study objectives | Discussion, par. 1:<br>"Using a MR approach, we present evidence for the involvement of placenta in susceptibility to broadly defined depression. Specifically, we ..." |
| 15 | **Limitations** | Discuss limitations of the study, taking into account the validity of the IV assumptions, other sources of potential bias, and imprecision. Discuss both direction and magnitude of any potential bias and any efforts to address them | Discussion, par. 5:<br>"The findings in this work have to be interpreted in the context of a MR approach. The reliability of MR results is based on several IV assumptions. While the …"<br>Discussion, par. 6:<br>"Another limitation was the low sample size of some of the GWAS of psychiatric disorders reducing the power of the analysis. This is especially |

| | | | |
|---|---|---|---|
| | | | problematic in ...” |
| 16 | **Interpretation** | | |
| | a) | Meaning: Give a cautious overall interpretation of results in the context of their limitations and in comparison, with other studies | Discussion, par. 2:<br>“A previous MR study analyzed effects of birth weight on several psychiatric disorders, using SNPs with direct fetal effect on birth weight as IVs [17]. In contrast with our results, the authors …”<br>Discussion, par. 3:<br>“The outcome GWAS used for depression was based on a broad, highly heterogeneous, phenotype, which includes from people with self-reported depressive symptoms to patients ascertained with a diagnostic interview [...] Thus, it may be possible that the causal association we detected is not specific to depression, but it was the only significant result in MR due to higher power. This agrees with the epidemiological study of Pettersson et al. [11], who found an association of birth weight with a general psychopathology factor.”<br>Discussion, par. 5:<br>“The findings in this work have to be interpreted in the context of a MR approach...” |
| | b) | Mechanism: Discuss underlying biological mechanisms that could drive a potential causal relationship between the investigated exposure and the outcome, and whether the gene-environment equivalence assumption is reasonable. Use causal language carefully, clarifying that IV estimates may provide causal effects only under certain assumptions | Discussion, par. 4:<br>“The causal effect of trophoblast functioning on susceptibility to psychopathology may be related to the process of invasion of the uterine walls that takes place in the early stages of placentation. The aim of this process is …”<br>Discussion, par. 5:<br>“The findings in this work have to be interpreted in the context of a MR approach. The reliability of MR results is based on several IV assumptions. […] However, we cannot discard an effect of the selected IVs by another mechanism not related with fetal growth restriction.” |
| | c) | Clinical relevance: Discuss whether the results have clinical or public policy relevance, and to what extent they inform effect sizes of possible interventions | Discussion, par. 7:<br>“Although no treatments for fetal growth restriction are currently available, there are several promising agents on clinical trials or preclinical research, acting on processes such as oxygen supply by angiogenesis or vasodilatation, or mitigation of oxidative stress. Therefore, further research is needed to confirm this causal effect, as it could have clear implications |

| | | | |
|---|---|---|---|
| | | | in prevention of mental disorders in near future.” |
| 17 | **Generalizability** | Discuss the generalizability of the study results (a) to other populations, (b) across other exposure periods/timings, and (c) across other levels of exposure | Discussion, par. 6:<br>“Finally, the studies included in the PlacentalCellEnrich database use expression data for placentas in the first three months of pregnancy. Although this period is key for establishment of the maternal-fetal interface, it would be necessary to include results from placentas in the second and third trimester to obtain a more complete view of placenta-associated variants.” |
| **OTHER INFORMATION** | | | |
| 18 | **Funding** | Describe sources of funding and the role of funders in the present study and, if applicable, sources of funding for the databases and original study or studies on which the present study is based | Section “Financial support” |
| 19 | **Data and data sharing** | Provide the data used to perform all analyses or report where and how the data can be accessed, and reference these sources in the article. Provide the statistical code needed to reproduce the results in the article, or report whether the code is publicly accessible and if so, where | Supplementary Table 2 |
| 20 | **Conflicts of interests** | All authors should declare all potential conflicts of interest | Section “Competing interests” |

Supplementary Table 2. Descriptive information on the GWAS used in the analysis.

| Trait | N | N cases | N controls | Mean (s.d.) | Diagnostic criteria[a] | Reference |
|---|---|---|---|---|---|---|
| **Birthweight** | 423,683 | | | 3,715 g (492) | Exclusion of multiple births, infant deaths and out-of-term births (gestational age <258 d and gestational age ≥294 d). Any extreme outliers in the birth weight. Babies born with congenital anomalies. UKBB only included individuals between 2.5 and 4.5 kg. | Juliusdottir et al. (5) |
| **Alcohol dependence** | 38,686 | 10,206 | 28,480 | | Diagnose by DSM-IV (or DSM-IIIR) criteria derived either from clinician ratings or semistructured interviews. | Walters et al. (6) |
| **Attention-deficit/ hyperactivity disorder** | 225,534 | 38,691 | 186,843 | | Singletons diagnosed by psychiatrists according to the ICD10 criteria (F90.0, F90.1, F98.8 diagnosis codes) or individuals who had been prescribed medication specific for ADHD symptoms (ATC-NA06BA, mostly methylphenidate). | Demontis et al. (7) |
| **Autism spectrum disorder** | 46,350 | 18,381 | 27,969 | | Singletons diagnosed with ASD by a psychiatrist according to ICD10, including diagnoses of childhood autism (ICD10 code F84.0), atypical autism (F84.1), Asperger's syndrome (F84.5), other pervasive developmental disorders (F84.8), and pervasive | Grove et al. (8) |

| | | | | | |
|---|---|---|---|---|---|
| | | | | developmental disorder unspecified (F84.9). | |
| **Bipolar disorder** | 413,466 | 41,917 | 371,549 | Required to meet international consensus criteria (DSM-IV, ICD-9 or ICD-10) for a lifetime diagnosis of BD, established using structured diagnostic instruments from assessments by trained interviewers, clinician-administered checklists or medical record review. | Mullins et al. (9) |
| **Cannabis use disorder** | 357,806 | 14,080 | 343,726 | PGC cases met criteria for a lifetime diagnosis of DSM-IV (or DSM-III-R) cannabis abuse or dependence derived from clinician ratings or semi-structured interviews. iPSYCH sample had ICD-10 codes of F12.1 (cannabis abuse) or F12.2 (cannabis dependence), or both. deCODE sample met criteria for lifetime DSM-III-R or DSM-IV cannabis abuse or dependence or DSM-5 cannabis use disorder. | Johnson et al. (10) |
| **Major depressive disorder** | 500,199 | 170,756 | 329,443 | Three depression phenotypes were assessed ranged self-reported help-seeking for problems with nerves, anxiety, tension or depression (termed ‘broad depression’), probable MDD based on self-reported depressive symptoms with associated impairment, and MDD identified from hospital | Howard et al. (11) |

| | | | | | | |
|---|---|---|---|---|---|---|
| | | | | | admission records. Exclusions were applied to participants who were identified with bipolar disorder, schizophrenia, or personality disorder using self-declared data as well as prescriptions for antipsychotic medications. | |
| **Post-traumatic stress disorder** | 174,659 | 23,212 | 151,447 | | Diagnose was based either on lifetime (where possible) or current PTSD (i.e., including participants with a potential lifetime PTSD diagnosis as controls), and PTSD diagnosis was established using various instruments and different versions of the DSM (DSM-III-R, DSM-IV, DSM-5). | Nievergelt et al. (12) |
| **Schizophrenia** | 130,644 | 53,386 | 77,258 | | Consensus diagnosis (DSM-IV, ICD-10), research diagnostic interview, review of medical records, mixed strategy. | Trubetskoy et al. (13) |

[a]For further details see the original paper (Reference column).

Supplementary Table 3. Instrumental variables used in the MR analysis associated to GO term "embryonic placenta development".

| SNP ID | Position hg38 | Alleles[a] | EAF[b] | Beta (s.e.) birth weight | P birth weight | Gene in GO term | $R^2$ | F-statistic |
|---|---|---|---|---|---|---|---|---|
| rs1374204 | Chr2:46257066 | C/T | 0.2813 | -0.0472(0.0028) | 4.4E-64 | *EPAS1* | 9.24E-04 | 391.97 |
| rs4953353 | Chr2:46340137 | T/G | 0.3698 | -0.0192(0.0026) | 9.7E-14 | *EPAS1* | 1.71E-04 | 72.49 |
| rs577204588[c] | Chr6:53156939 | C/T | 0.0010 | -0.1535(0.0234) | 5.8E-11 | *GCM1* | 2.48E-04 | 105.09 |
| rs11042596 | Chr11:2097630 | T/G | 0.3807 | 0.026(0.0027) | 1.3E-22 | *IGF2* | 2.96E-04 | 125.56 |
| rs1011476[c] | Chr11:2277805 | T/G | 0.2465 | 0.0179(0.0027) | 4E-11 | *ASCL2, IGF2* | 1.30E-04 | 55.15 |
| rs234864[c] | Chr11:2836067 | G/A | 0.4523 | -0.0169(0.0025) | 1.2E-11 | *CDKN1C* | 1.42E-04 | 60.22 |
| rs4444073[c] | Chr11:10310117 | C/A | 0.4732 | -0.0219(0.0024) | 2.8E-19 | *ADM* | 2.39E-04 | 101.39 |
| rs55836809 | Chr13:27928737 | G/A | 0.2316 | -0.0204(0.003) | 1.9E-11 | *CDX2* | 1.41E-04 | 59.87 |
| rs55958435 | Chr15:96309409 | G/A | 0.2127 | -0.0247(0.0028) | 4.3E-18 | *NR2F2* | 2.32E-04 | 98.31 |
| rs41355649[c] | Chr19:33299650 | A/G | 0.0646 | -0.0366(0.0051) | 5E-13 | *CEBPA* | 1.63E-04 | 69.09 |
| rs1135856 | Chr22:46044725 | C/T | 0.2684 | -0.0192(0.0028) | 5.6E-12 | *WNT7B* | 1.51E-04 | 63.81 |

[a]First allele is the effect allele

[b]Effect allele frequency in the 1000Genomes reference panel (European population)

[c]Common to main analysis.

#### 2 References

1. Jain A, Tuteja G. PlacentaCellEnrich: A tool to characterize gene sets using placenta cell-specific gene enrichment analysis. Placenta (2021) 103:164–171. doi: 10.1016/J.PLACENTA.2020.10.029
